# Analysis of Costs and Diagnostic Experience of Stroke Mimics at a Latin American University Hospital

**DOI:** 10.1101/2024.05.28.24308086

**Authors:** Juan David Villalobos Ibarra, Jorge Carrizosa, Andrea Vargas, Carlos Martinez, Denise Battaglini, Daniel Agustin Godoy

**Affiliations:** Department of critical and intensive care medicine, academic hospital Fundación Santa Fe de Bogotá, Bogotá, Colombia. Universidad del Rosario, School of Medicine, Program of Critical and Intensive Care, Bogotá, Colombia; Neurology Department, academic Hospital Fundación Santa Fe de Bogotá, Bogotá, Colombia; UO Clinica Anestesiologica e Terapia Intensiva, IRCCS Ospedale Policlinico San Martino, Genova, Italia; Neurointensive Care Unit, Sanatorio Pasteur, Catamarca, Argentina

## Abstract

**Introduction:** The incidence of stroke mimic symptoms is around 30%. These symptoms impact healthcare costs, often leading mimic stroke patients to undergo unnecessary clinical tests, imaging, and treatments.

**Objectives:** The main goal of this study is to describe the types of stroke mimic symptoms and estimate healthcare costs in patients with stroke mimics. Secondary objectives include comparing costs based on Telestroke Mimics Stroke and FABS scores and determining the frequency of thrombolysis.

**Methods:** We conducted a retrospective observational study. We reviewed medical records of all patients admitted to Fundación Santa Fe de Bogotá with a final diagnosis of ischemic stroke mimic. We characterized the study population and analyzed the costs of interventions in these patients.

**Results:** A total of 111 patients were included. The average age at mimic presentation was 65 ± 19.4 years, with transient ischemic attack (TIA) being the most common cause of mimics in both sexes, followed by migraine. Tissue plasminogen activator was administered in 0.9% of patients. The direct costs of activating the stroke code averaged US$1,098.72, with a cost of laboratory and imaging at US$773.95. The average cost of total hospitalization was US$2,220.16 per patient.

**Conclusions:** The most frequent cause of stroke mimic was TIA, and thrombolysis was performed in 0.9% of cases. The direct costs incurred by activating the stroke code, diagnostic tests, treatment with intravenous thrombolysis, and hospitalization of patients with stroke mimic are lower compared to other studies.

## Introduction

A stroke mimic is a non-vascular condition that causes symptoms similar to those of an acute stroke (1). These patients often end up in the emergency room, with 30% of them showing signs and symptoms of stroke (2).

A stroke mimic is considered when a diagnosis of acute ischemic stroke has been ruled out, and another cause for the initial symptoms is identified (3). Common causes of stroke mimics include epilepsy, syncope, sepsis, psychiatric disorders, migraine, brain tumors, metabolic diseases, neuropathies, and other neurovascular disorders (4).

The cost of ischemic stroke in the United States between 2017 and 2018 was nearly 53 billion US dollars, including healthcare services, medications, and lost productivity (5). Direct and indirect costs of managing a stroke mimic in four US centers were found to be 355,198 US dollars and 288,277 US dollars, respectively, with an average cost of 5,401 US dollars per hospital admission (6). A study conducted in the Philippines at a tertiary hospital accredited by the Joint Commission International (JCI) revealed that the cost of laboratory and imaging tests performed from stroke code activation until an alternative diagnosis is made is approximately 500 US dollars (7).

Misdiagnosing stroke mimics leads to unnecessary tests and treatments, increasing the risk of complications and costs for patients. Prompt recognition of stroke mimics can reduce the burden of healthcare costs by avoiding unnecessary clinical tests, imaging, and treatments (8).

## Methods

The main goal of this study was to describe the symptoms of stroke mimics and assess healthcare costs in these patients. Secondary goals included comparing costs based on Telestroke Mimic and FABS scores and determining the frequency of thrombolysis.

From 2017 to 2020, we conducted a descriptive retrospective observational study at Fundación Santa Fe de Bogotá, Colombia on patients over 18 years old who were initially thought to have symptoms of a stroke but were later considered to have stroke mimics. We excluded patients with ischemic stroke, those under 18 years old, those referred from other institutions more than 6 hours after symptoms started, and those referred to another institution before the diagnosis of stroke mimic. The study followed the World Medical Association (WMA) Declaration of Helsinki – Ethical Principles for Medical Research involving Human Subjects and adhered to the STrengthening the Reporting of OBservational studies in Epidemiology (STROBE) guidelines.

Our protocol was approved by the scientific Committee and ethics committee of Fundación Santa Fe de Bogotá, Colombia and by Universidad del Rosario, Colombia (approval number CCEI-12641-2020). We collected demographic, clinical, imaging, and intravenous thrombolysis data from the stroke team registry and medical records at our institution. Direct hospitalization costs were obtained from the financial department and collaboration with the technology and information technology teams at Fundación Santa Fe de Bogotá.

At our institution, physicians, nurses, and laboratory/radiology personnel are trained to identify stroke symptoms, use scales, and follow a protocol to activate a stroke code, request laboratories, and neuroimaging. A neurology specialist evaluates the patient, and the decision to perform additional exams, confirm a stroke, identify a stroke mimic, or administer rt-PA is made following stroke guidelines.

We did not perform a sample size calculation for this exploratory study. Qualitative variables were expressed as proportions or percentages, quantitative variables as averages and standard deviations (SD) or medians and interquartile range (IQR) depending on their distribution. Categorical variables were described in terms of percentages. Costs were summarized by means of medians and IQR in dollars, considering the World Bank conversion rate for the corresponding year, globally and stratified by treatment (thrombolysis as a dichotomous variable). All analyses were conducted using SPSS v.22 and R software.

## Results

In the study, 144 patients were screened for inclusion. 33 patients were excluded due to not meeting the inclusion criteria or due to lack of data for analysis. Finally, 111 patients who met the inclusion criteria were studied. The median age of presentation with mimic was 68.0 years (ranging from 50.0 to 82.0), and 54.1% of the patients were female. The most common comorbidities, in order of frequency, were arterial hypertension, diabetes mellitus, dyslipidemia, cancer, previous coronary syndrome, and migraine. The most frequently used medications at home were antihypertensives (41.4%) and antiplatelet drugs or anticoagulants (20.7%) (Table 1).

**Table 1.**
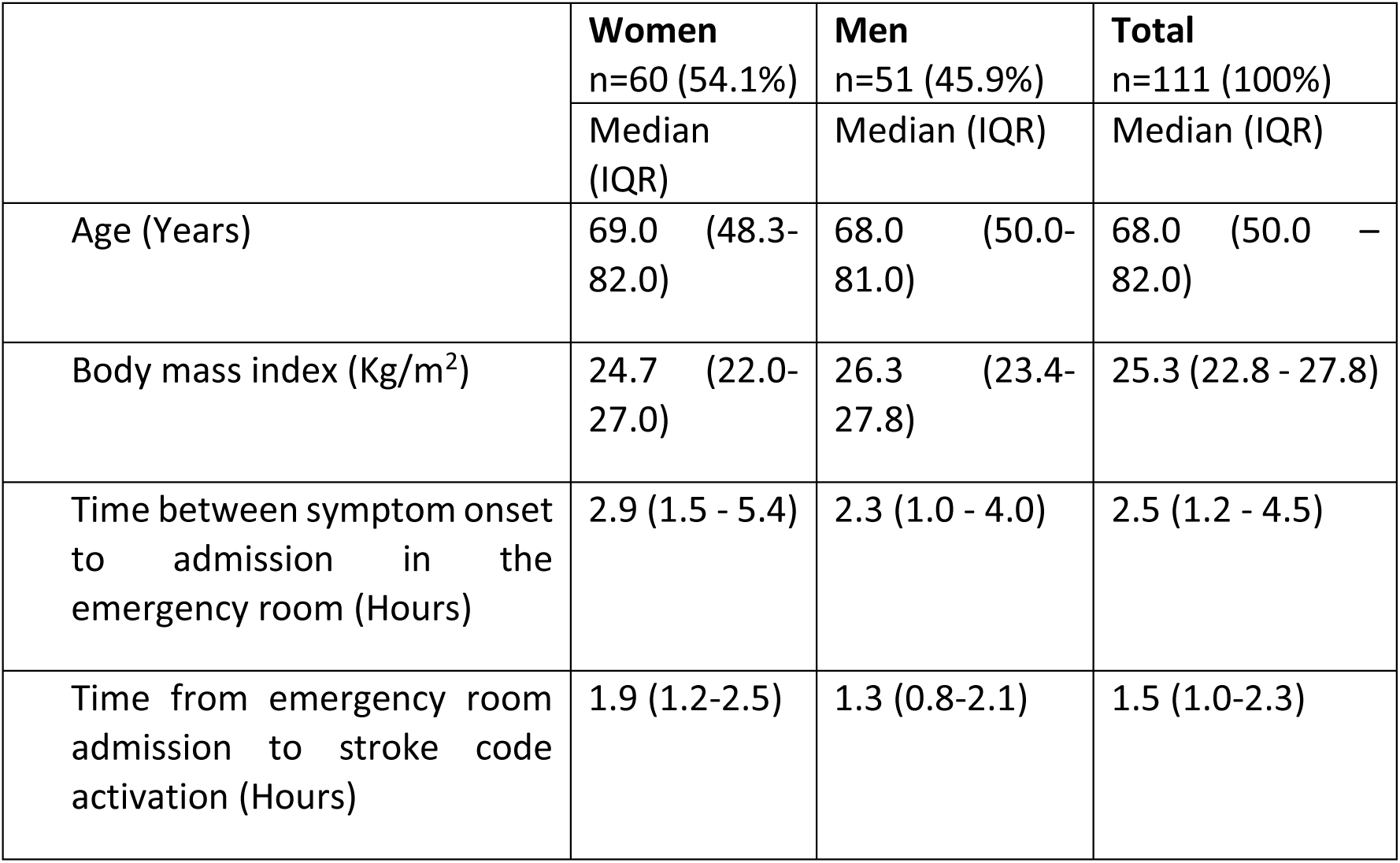
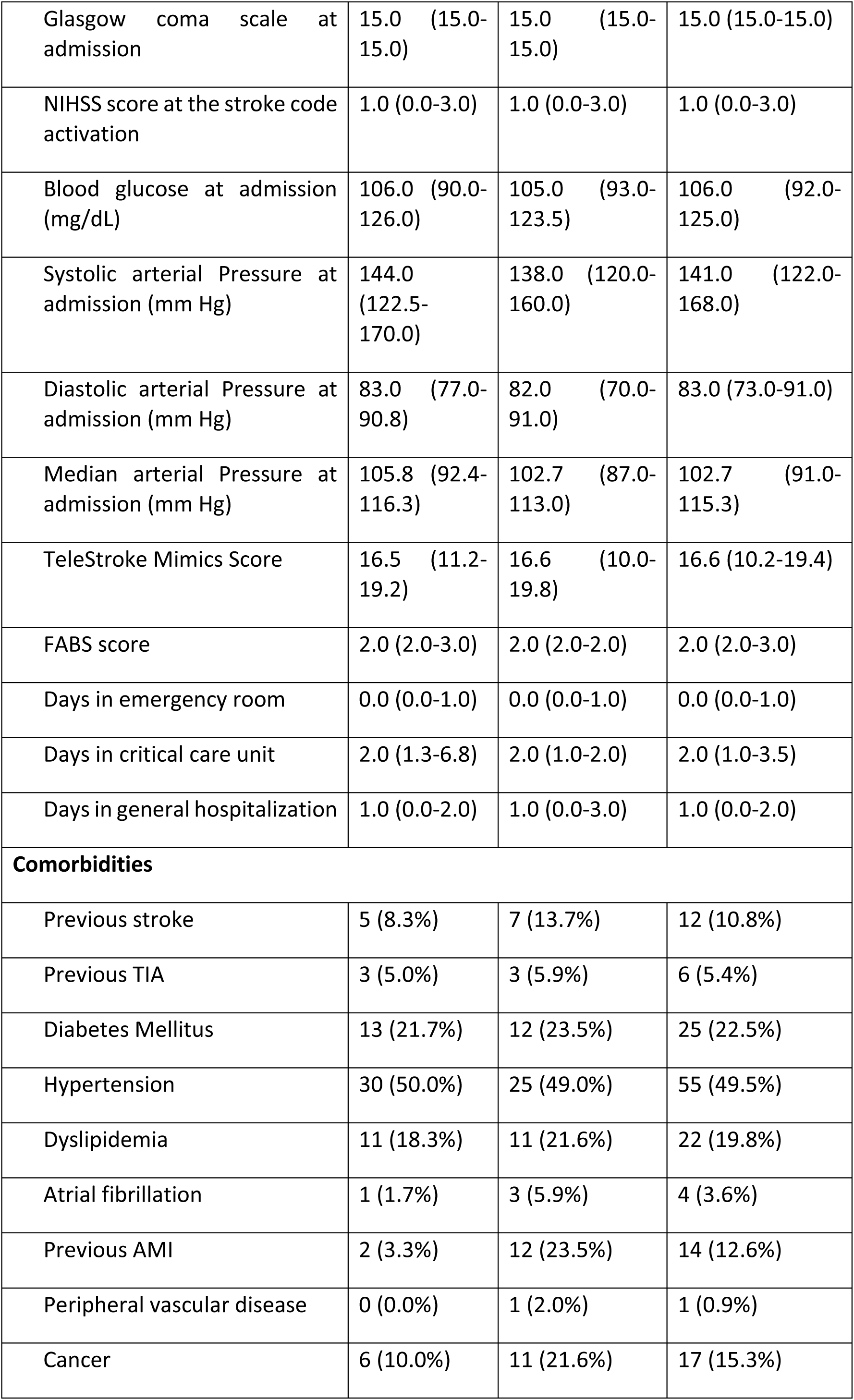
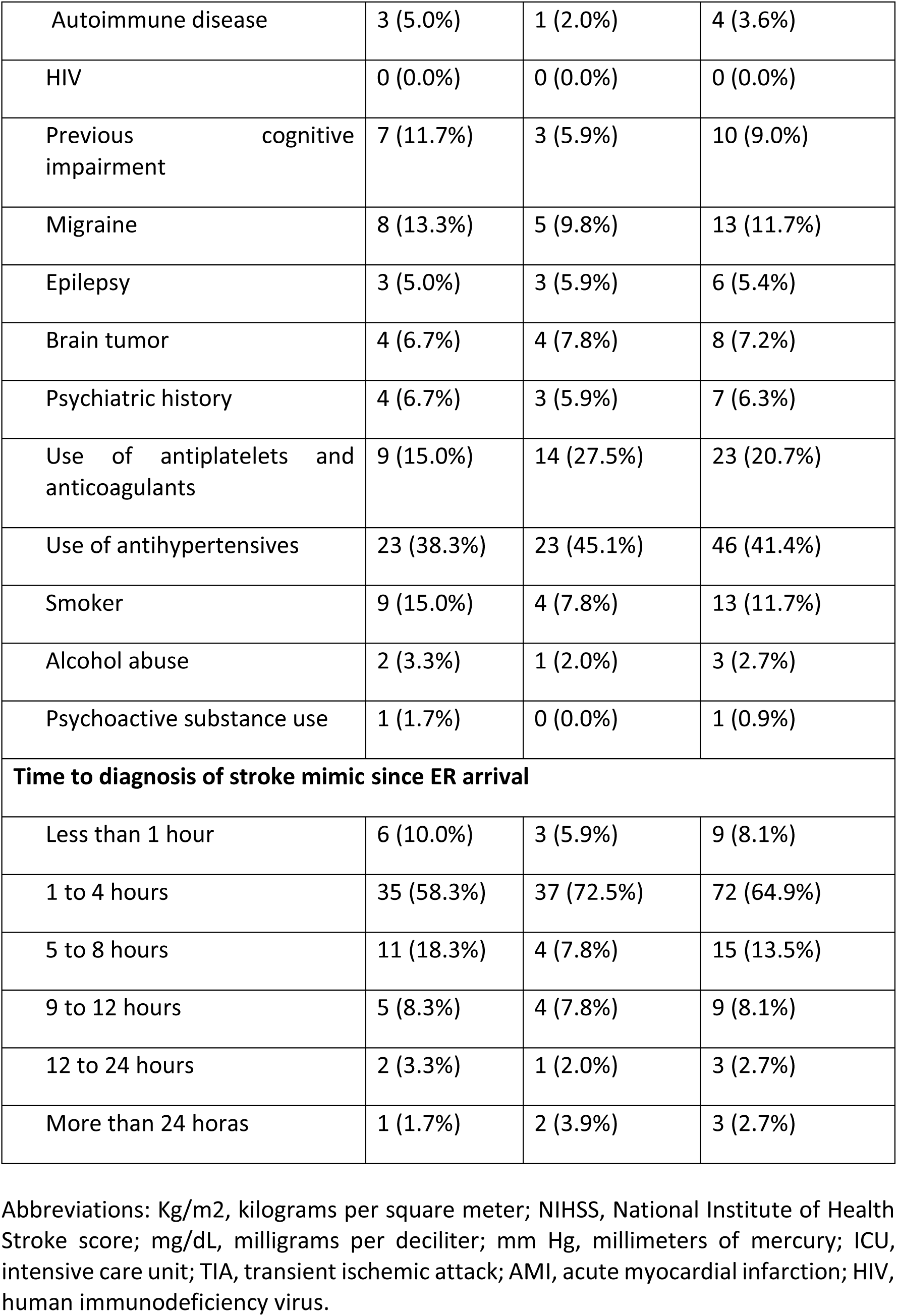
Characteristics of the patients according to the gender.

|  | **Women**<br>n=60 (54.1%) | **Men**<br>n=51 (45.9%) | **Total**<br>n=111 (100%) |
| --- | --- | --- | --- |
|  | Median<br>(IQR) | Median (IQR) | Median (IQR) |
| Age (Years) | 69.0 (48.3-82.0) | 68.0 (50.0-81.0) | 68.0 (50.0 – 82.0) |
| Body mass index (Kg/m<sup>2</sup>) | 24.7 (22.0-27.0) | 26.3 (23.4-27.8) | 25.3 (22.8 - 27.8) |
| Time between symptom onset to admission in the emergency room (Hours) | 2.9 (1.5 - 5.4) | 2.3 (1.0 - 4.0) | 2.5 (1.2 - 4.5) |
| Time from emergency room admission to stroke code activation (Hours) | 1.9 (1.2-2.5) | 1.3 (0.8-2.1) | 1.5 (1.0-2.3) |
| Glasgow coma scale at admission | 15.0 (15.0-15.0) | 15.0 (15.0-15.0) | 15.0 (15.0-15.0) |
| NIHSS score at the stroke code activation | 1.0 (0.0-3.0) | 1.0 (0.0-3.0) | 1.0 (0.0-3.0) |
| Blood glucose at admission (mg/dL) | 106.0 (90.0-126.0) | 105.0 (93.0-123.5) | 106.0 (92.0-125.0) |
| Systolic arterial Pressure at admission (mm Hg) | 144.0 (122.5-170.0) | 138.0 (120.0-160.0) | 141.0 (122.0-168.0) |
| Diastolic arterial Pressure at admission (mm Hg) | 83.0 (77.0-90.8) | 82.0 (70.0-91.0) | 83.0 (73.0-91.0) |
| Median arterial Pressure at admission (mm Hg) | 105.8 (92.4-116.3) | 102.7 (87.0-113.0) | 102.7 (91.0-115.3) |
| TeleStroke Mimics Score | 16.5 (11.2-19.2) | 16.6 (10.0-19.8) | 16.6 (10.2-19.4) |
| FABS score | 2.0 (2.0-3.0) | 2.0 (2.0-2.0) | 2.0 (2.0-3.0) |
| Days in emergency room | 0.0 (0.0-1.0) | 0.0 (0.0-1.0) | 0.0 (0.0-1.0) |
| Days in critical care unit | 2.0 (1.3-6.8) | 2.0 (1.0-2.0) | 2.0 (1.0-3.5) |
| Days in general hospitalization | 1.0 (0.0-2.0) | 1.0 (0.0-3.0) | 1.0 (0.0-2.0) |
| **Comorbidities** |  |  |  |
| Previous stroke | 5 (8.3%) | 7 (13.7%) | 12 (10.8%) |
| Previous TIA | 3 (5.0%) | 3 (5.9%) | 6 (5.4%) |
| Diabetes Mellitus | 13 (21.7%) | 12 (23.5%) | 25 (22.5%) |
| Hypertension | 30 (50.0%) | 25 (49.0%) | 55 (49.5%) |
| Dyslipidemia | 11 (18.3%) | 11 (21.6%) | 22 (19.8%) |
| Atrial fibrillation | 1 (1.7%) | 3 (5.9%) | 4 (3.6%) |
| Previous AMI | 2 (3.3%) | 12 (23.5%) | 14 (12.6%) |
| Peripheral vascular disease | 0 (0.0%) | 1 (2.0%) | 1 (0.9%) |
| Cancer | 6 (10.0%) | 11 (21.6%) | 17 (15.3%) |
| Autoimmune disease | 3 (5.0%) | 1 (2.0%) | 4 (3.6%) |
| HIV | 0 (0.0%) | 0 (0.0%) | 0 (0.0%) |
| Previous cognitive impairment | 7 (11.7%) | 3 (5.9%) | 10 (9.0%) |
| Migraine | 8 (13.3%) | 5 (9.8%) | 13 (11.7%) |
| Epilepsy | 3 (5.0%) | 3 (5.9%) | 6 (5.4%) |
| Brain tumor | 4 (6.7%) | 4 (7.8%) | 8 (7.2%) |
| Psychiatric history | 4 (6.7%) | 3 (5.9%) | 7 (6.3%) |
| Use of antiplatelets and anticoagulants | 9 (15.0%) | 14 (27.5%) | 23 (20.7%) |
| Use of antihypertensives | 23 (38.3%) | 23 (45.1%) | 46 (41.4%) |
| Smoker | 9 (15.0%) | 4 (7.8%) | 13 (11.7%) |
| Alcohol abuse | 2 (3.3%) | 1 (2.0%) | 3 (2.7%) |
| Psychoactive substance use | 1 (1.7%) | 0 (0.0%) | 1 (0.9%) |
| **Time to diagnosis of stroke mimic since ER arrival** |  |  |  |
| Less than 1 hour | 6 (10.0%) | 3 (5.9%) | 9 (8.1%) |
| 1 to 4 hours | 35 (58.3%) | 37 (72.5%) | 72 (64.9%) |
| 5 to 8 hours | 11 (18.3%) | 4 (7.8%) | 15 (13.5%) |
| 9 to 12 hours | 5 (8.3%) | 4 (7.8%) | 9 (8.1%) |
| 12 to 24 hours | 2 (3.3%) | 1 (2.0%) | 3 (2.7%) |
| More than 24 horas | 1 (1.7%) | 2 (3.9%) | 3 (2.7%) |

Abbreviations: Kg/m<sup>2</sup>, kilograms per square meter; NIHSS, National Institute of Health Stroke score; mg/dL, milligrams per deciliter; mm Hg, millimeters of mercury; ICU, intensive care unit; TIA, transient ischemic attack; AMI, acute myocardial infarction; HIV, human immunodeficiency virus.

The median time from symptom onset to admission to the emergency room was 2.5 hours (ranging from 1.2 to 4.5 hours). The median time for stroke code activation was 1.5 hours (ranging from 1.0 to 2.3 hours), with a median NIHSS score at code activation of 1 point. The median Telestroke Mimics Stoke and FABS scores were 16.6 (ranging from 10.2 to 19.4) and 2.0 (ranging from 2.0 to 3.0) respectively. The median length of stay in the emergency room was 0.0 days (ranging from 0.0 to 1.0 days), in the intensive care unit was 2.0 days (ranging from 1.0 to 3.5 days), and in general hospitalization was 1.0 days (ranging from 0.0 to 2.0 days) before discharge (Table 1).

The time required from admission at the emergency room to the diagnosis of stroke mimic was between 1 and 4 hours in 64.9% of the patients (Table 1). Brain magnetic resonance imaging (MRI) was the first neuroimaging performed in 57.7% of the patients, followed by head computed tomography (CT) in 36.9% of the patients. Neuroimaging results were normal in 93.7% of the patients. An intracranial space-occupying lesion was found in 6.3% of the patients, with no presence of intracranial bleeding or cerebral ischemia in any of the patients (Table 2). Other diagnostic tests by frequency were: 27% transthoracic echocardiogram, 18% carotid Doppler ultrasound, 3.6% transesophageal echocardiogram, and 0.9% electrocardiographic Holter monitoring (Table 2).

**Table 2.** Neuroimaging and findings according to gender.

| First Neuroimaging in the | Women | Men | Total |
| --- | --- | --- | --- |
| **emergency room** | n=60 (54.1%) | n=51 (45.9%) | n=111 (100%) |
| No imaging performed | 3 (5.0%) | 2 (3.9%) | 5 (4.5%) |
| Head CT | 18 (30.0%) | 23 (45.1%) | 41 (36.9%) |
| Brain MRI | 39 (65.0%) | 25 (49.0%) | 64 (57.7%) |
| Brain MRA | 0 (0.0%) | 0 (0.0%) | 0 (0.0%) |
| Cerebral CT Angiography | 0 (0.0%) | 1 (2.0%) | 1 (0.9%) |
| **Findings in the first neuroimaging** |  |  |  |
| Normal | 58 (96.7%) | 46 (90.2%) | 104 (93.7%) |
| Cerebral ischemia | 0 (0.0%) | 0 (0.0%) | 0 (0.0%) |
| Intracranial hemorrhagic lesion | 0 (0.0%) | 0 (0.0%) | 0 (0.0%) |
| Intracranial space-occupying lesion | 2 (3.3%) | 5 (9.8%) | 7 (6.3%) |
| **Another diagnostic test performed** |  |  |  |
| Carotid Doppler ultrasound | 9 (15.0%) | 11 (21.6%) | 20 (18.0%) |
| Transthoracic echocardiogram | 13 (21.7%) | 17 (33.3%) | 30 (27.0%) |
| Transesophageal echocardiogram | 1 (1.7%) | 3 (5.9%) | 4 (3.6%) |
| Electrocardiographic Holter monitoring | 1 (1.7%) | 0 (0.0%) | 1 (0.9%) |

Abbreviations: CT, Computed tomography; MRI, Magnetic resonance imaging; MRA, Magnetic resonance angiography.

The most common etiology of stroke mimic in both sexes was TIA (18.9%) followed by migraine (15.3%), and epileptic seizures (9.9%). Administration of tissue plasminogen activator was performed in 1 (0.9%) patient, followed by bleeding complication in that patient (Table 3).

**Table 3.** Stroke mimic etiology. rt-PA administration and complications according to the gender.

| **Etiology** | **Women**<br>n=60 (54.1%) | **Men**<br>n=51 (45.9%) | **Total**<br>n=111 (100%) |
| --- | --- | --- | --- |
| Epileptic seizures | 4 (6.7%) | 7 (13.7%) | 11 (9.9%) |
| Central or peripheral neuropathy | 3 (5.0%) | 4 (7.8%) | 7 (6.3%) |
| Acute confusional state | 1 (1.7%) | 1 (2.0%) | 2 (1.8%) |
| Intracranial tumor | 1 (1.7%) | 3 (5.9%) | 4 (3.6%) |
| Peripheral vertigo | 3 (5.0%) | 2 (3.9%) | 5 (4.5%) |
| Migraine | 11 (18.3%) | 6 (11.8%) | 17 (15.3%) |
| Hypoglycemia | 1 (1.7%) | 0 (0.0%) | 1 (0.9%) |
| Hydroelectrolyte disorder | 1 (1.7%) | 1 (2.0%) | 2 (1.8%) |
| Syncope | 0 (0.0%) | 3 (5.9%) | 3 (2.7%) |
| Pharmacological agents | 2 (3.3%) | 0 (0.0%) | 2 (1.8%) |
| Psychoactive substance use | 0 (0.0%) | 0 (0.0%) | 0 (0.0%) |
| Central nervous system infection | 1 (1.7%) | 0 (0.0%) | 1 (0.9%) |
| Demyelinating disease | 0 (0.0%) | 0 (0.0%) | 0 (0.0%) |
| Posterior reversible encephalopathy syndrome | 0 (0.0%) | 1 (2.0%) | 1 (0.9%) |
| Anxiety disorder | 2 (3.3%) | 4 (7.8%) | 6 (5.4%) |
| Conversion disorder | 7 (11.7%) | 0 (0.0%) | 7 (6.3%) |
| Unclear diagnosis | 3 (5.0%) | 0 (0.0%) | 3 (2.7%) |
| Transient global amnesia | 2 (3.3%) | 3 (5.9%) | 5 (4.5%) |
| Hypertensive encephalopathy | 5 (8.3%) | 1 (2.0%) | 6 (5.4%) |
| Delirium | 2 (3.3%) | 3 (5.9%) | 5 (4.5%) |
| Transient ischemic attack (TIA) | 10 (16.7%) | 11 (21.6%) | 21 (18.9%) |
| Facial paralysis | 1 (1.7%) | 0 (0.0%) | 1 (0.9%) |
| **Fibrinolysis administration** |  |  |  |
| Recombinant tissue plasminogen activator administration (rt-PA) | 0 (0.0%) | 1 (2.0%) | 1 (0.9%) |
| Bleeding (complication) | 0 (0.0%) | 1 (2.0%) | 1 (0.9%) |

Direct costs were obtained in 110 patients of the 111 patients registered in the database. The costs are reported in US dollars, with an average total cost hospitalization of 2220.16 US (95% CI 1396.71-3043.61), with a standard deviation of 4337.19 US, median of 938 US, with a lowest cost of 28 US to a maximum cost of 33,549 US, with a non-normal distribution due to extreme data with most costs below than expected for the total hospitalization (Figure 1).

**Figure 1.**
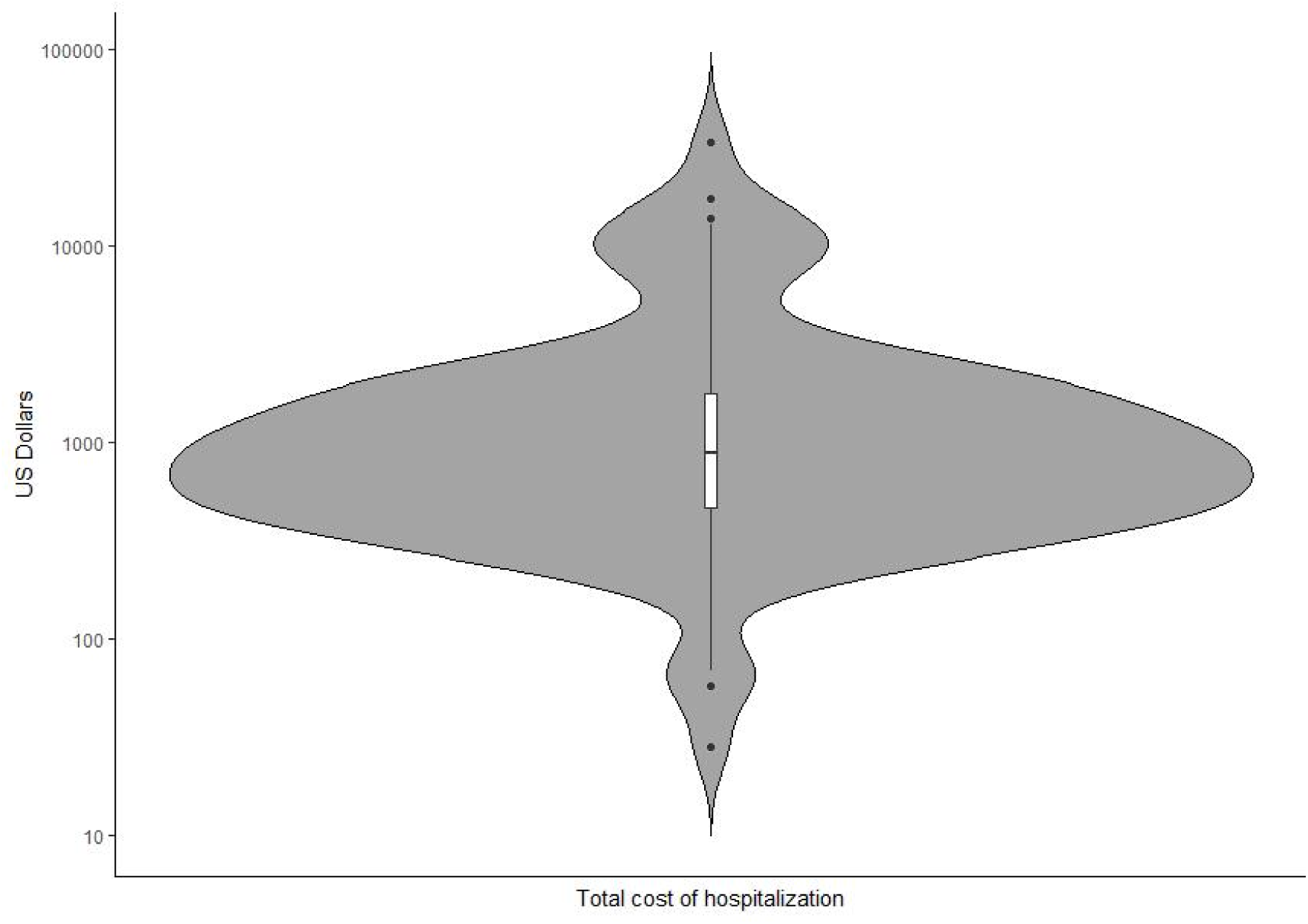
Average cost of hospitalization (US dollars)

Direct costs from activation to deactivation of the stroke code in 110/111 patients were on average 795.85 US (95% CI 589.18-1002.52 US), with a standard deviation of 1098.72 US, median of 511 US, with a lowest cost of 0 US to a maximum cost of 7721 US (Figure 2). The cost of the medication used during the activation code was on average of 21.9 US (95% CI 9.52-34.31), with a standard deviation of 65.88 US, median of 1.80 US, a lowest cost of 0 and a maximum cost of 436.8 US (Figure 3). Direct cost of the laboratory tests and imaging carried out on average was 773.95 US (95% CI 573.56-974.33) with a standard deviation of 1065.29 US, a lowest cost of 0 US and a maximum of 7467.93 US, with a non-normal distribution for the activation and deactivation of the stroke code due to extreme data, with most costs below than expected.

**Figure 2.**
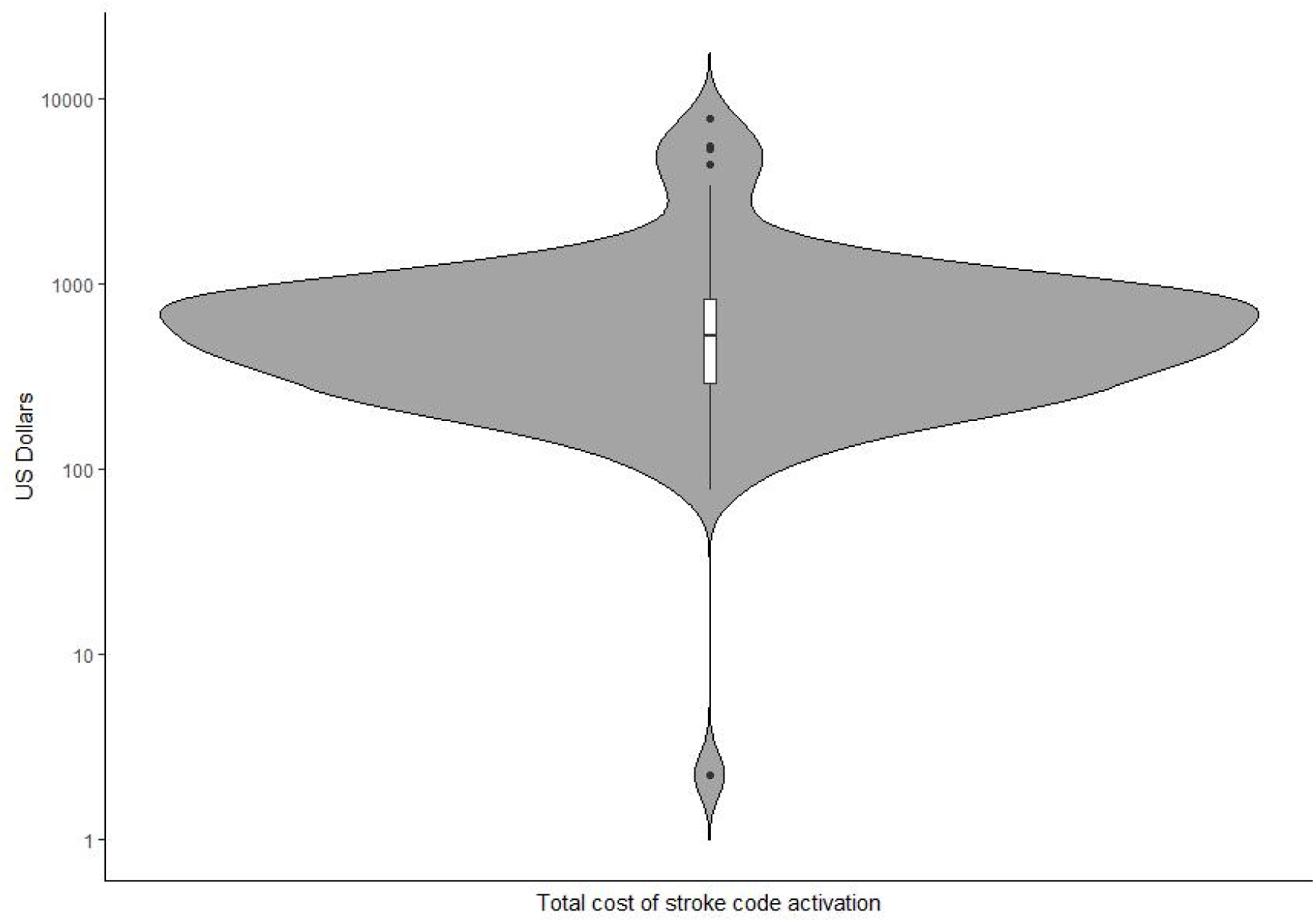
Average cost of stroke code activation (US dollars)

**Figure 3.**
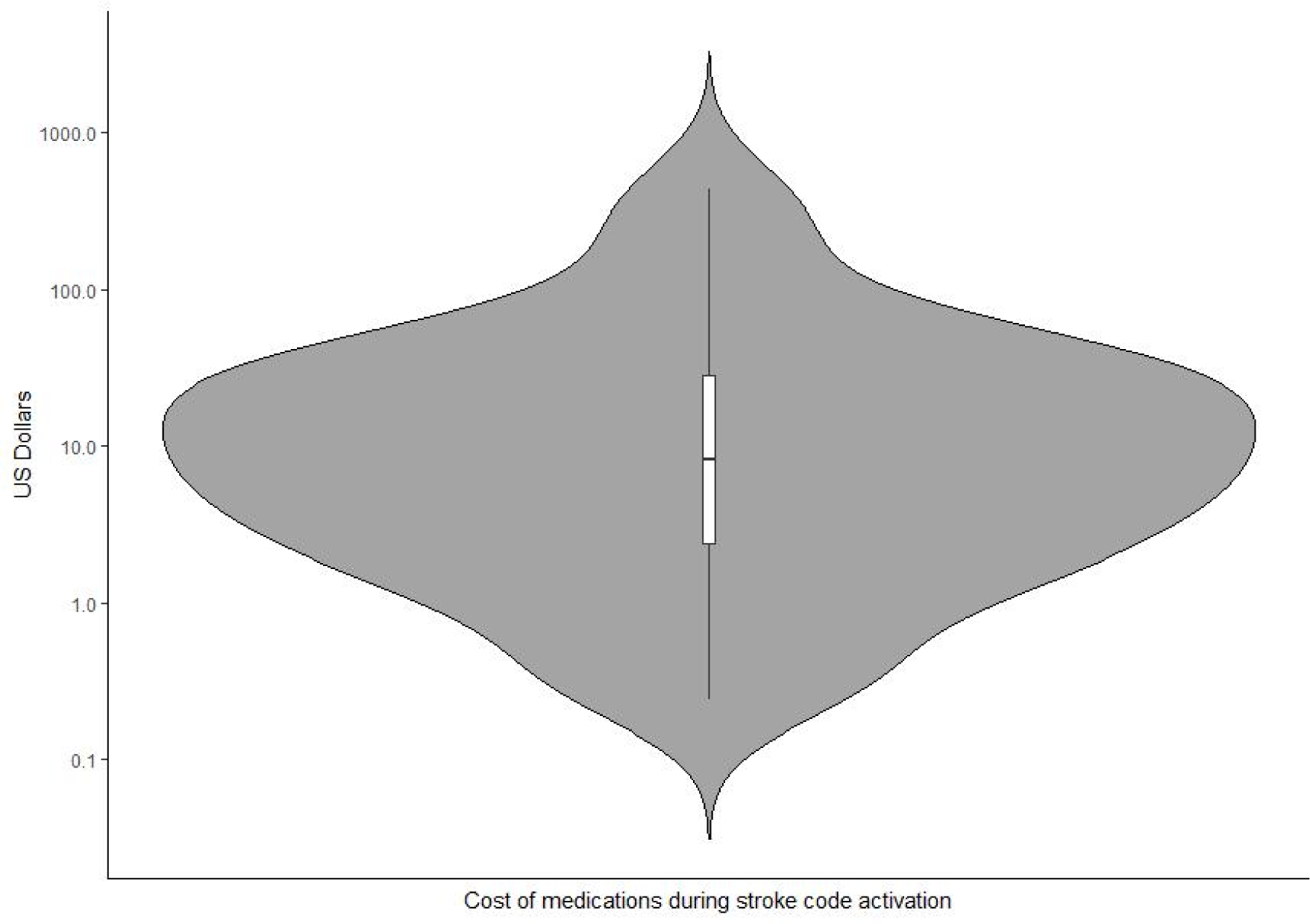
Average cost of medication used during stroke code activation (US dollars)

In the subgroup analysis, Telestroke score with more than 65% probability of being a mimic had an average cost of hospitalization of 2,294 US compared to an average cost of 1,786 US in those patients with Telestroke under 65% of probability (Figure 4). In addition, a FABS score ≥ 4 was obtained in 1/111 patients with an average cost of 9,071 US as compared to the average cost of hospitalization for patients with FABS score < 4 of 2,156 US.

**Figure 4.**
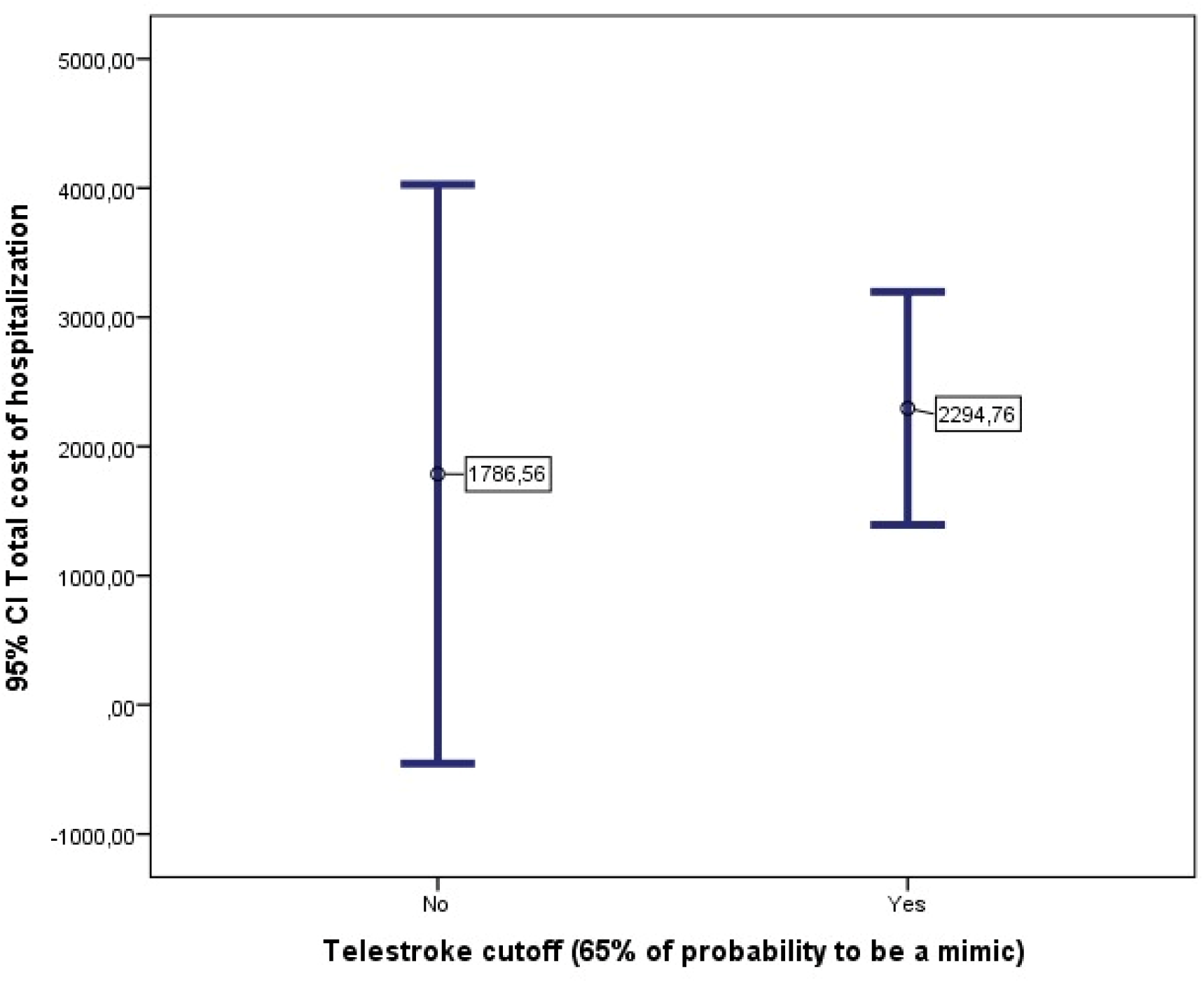
Average cost of hospitalization in patients with Telestroke score with 65% of probability VS under 65% of probability of being a stroke mimic (US dollars).

**Figure 5.**
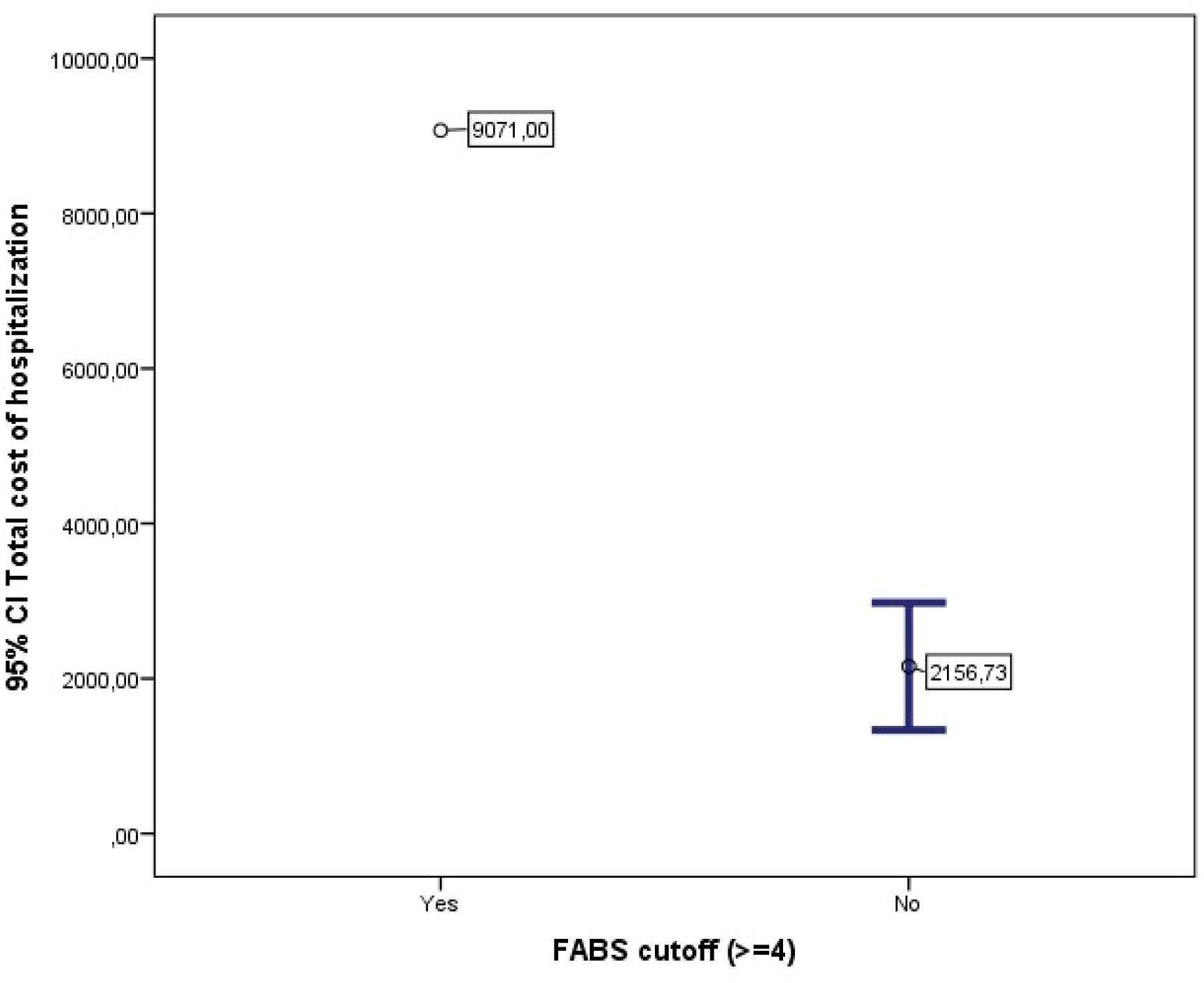
Average cost of hospitalization in patients with FABS score ≥ 4 (US dollars).

## Discussion

A stroke mimic is a non-vascular condition that presents as an acute neurological deficit clinically simulating acute ischemic stroke (1). Around 30% of patients admitted to the emergency room manifest signs and symptoms suggestive of stroke, despite being diagnosed as stroke mimics. This is more likely to occur in young people with mild symptoms and in the absence of risk factors (2), frequently presenting at admission with systolic blood pressures >140 mm Hg and NIHSS scores >5 (10).

In our study, patients with stroke mimic were on average 65 years old and predominantly female. The most common risk factors were arterial hypertension, diabetes mellitus, dyslipidemia, cancer, previous coronary syndrome, and migraine. Stroke mimic patients were admitted to the emergency department with an average systolic blood pressure of 146 mm Hg, which differs from other studies where stroke mimics are present in younger patients without risk factors (2) and systolic arterial pressures less than 140 mm Hg (10). We found a similar NIHSS scale of 3 points as reported by previous researchers as less than 5 points (10). Telestroke Mimic Stroke and FABS scores were associated with a high probability of stroke mimic (84.7% and 99.1% respectively).

In the present study, the most common causes of stroke mimic in both genders were: TIA, migraine, epileptic seizures, central or peripheral neuropathy, conversion disorder, anxiety, peripheral vertigo, hypertensive encephalopathy, transient amnesia, among others, which are very similar to the causes reported by previous studies (4).

As part of the ischemic stroke management, the use of intravenous plasminogen activator is recommended when stroke is suspected (11). Due to the limited time window period and the need to treat the suspected stroke, it is not uncommon for intravenous thrombolysis to be administered to patients who were true mimics, with a post-administration complication rate of 1%, with intracerebral hemorrhage being the most frequent complication (12). In our study, the administration of tissue plasminogen activator was performed in 0.9% of the patients who had stroke mimics, with a complication rate of 0.9% due to gastrointestinal bleeding. None of the patients in this study were later diagnosed as having a stroke. On the other hand, when evaluating the length of hospital stay of our patients, it was shorter than previous studies, with an average stay of 0.4 ± 0.7 days at the emergency room, 2.9 ± 2.4 days at the intensive care unit, and 2.1 ± 3.9 days at the hospital (13).

The cost of ischemic stroke in the United States between 2017 and 2018 was nearly 53 billion, including health care services, medications, and missed days of work (5). In the study by Goyal et. Al, direct and indirect cost of having a stroke mimic is US $355,198.0 and US $288,277.0 respectively (6). In the present study, the direct costs were analyzed, finding data on 110 of the 111 patients who met the inclusion criteria with a total average cost of hospitalization of US $2,220.16 per patient, with an average cost between the activation of the stroke code and its deactivation of US $795.85. It is observed that the costs per mimic in the Fundación Santa Fe de Bogotá were lower compared to other studies carried out in international institutions because they did more intravenous tissue plasminogen administration, neuroimaging and they analyze the cost of other therapies.

A study conducted in the Philippines at a tertiary hospital accredited by the Joint Commission International (JCI) between 2014 and 2017 found that the cost of laboratory and imaging performed from the patient’s stroke code activation until a stroke mimic was diagnosed was approximately US $500 (7), with a difference of US $273.95 when compared to the costs analyzed in the present study. However, in the mentioned study (7) it is not specified what type of neuroimaging and laboratories were performed to rule out stroke.

Tertiary hospitals are high-complexity facilities with specialized and subspecialized services (14). High-income countries spend about 100 times more on health per capita than low and middle-income countries where the resources for financing health services are limited (15). This may explain the differences in health costs between countries.

Additionally, in the subgroup analysis in patients with Telestroke Mimic Stroke with a 65% probability of being a mimic, the average cost is higher compared to those with Telestroke Mimic Stroke less than 65%. Additionally, the cost for patients with FABS score ≥4 was evaluated, however only one patient could be included within this subgroup, and it was not statistically significant.

## Conclusions

We have provided descriptive data from a leading stroke management center in Colombia. We found that stroke mimicry was more common in women and was linked to multiple other health conditions. Transient ischemic attack (TIA) was the most common initial presentation. Only 0.9% of patients with a stroke mimic received intravenous thrombolysis with tissue plasminogen activator before a stroke diagnosis was ruled out. Additionally, when we compared the direct costs associated with the stroke code activation, diagnostic tests, administration of intravenous thrombolysis, and hospitalization, we found that these costs were lower compared to similar studies conducted in international institutions. In our subgroup analysis, we observed that the costs were higher for patients with Telestroke scores suggesting a >65% probability of being a mimic, but we didn’t find significant cost differences for patients with a FABS score of ≥4.

## Data Availability

All data referred to in the manuscript is available for review/consult

## References

1. Vilela P. Acute stroke differential diagnosis: Stroke mimics. European Journal of Radiology. 2017.

2. Merino JG, Luby M, Benson RT, Davis LA, Hsia AW, Latour LL, et al. Predictors of acute stroke mimics in 8187 patients referred to a stroke service. Journal of Stroke and Cerebrovascular Diseases. 2013;

3. Anathhanam S, Hassan A. Mimics and chameleons in stroke. Clinical Medicine, Journal of the Royal College of Physicians of London. 2017;

4. Fernandes PM, Whiteley WN, Hart SR, Al-Shahi Salman R. Strokes: Mimics and chameleons. Pract Neurol. 2013;

5. Prevention C for disease control and. Stroke Facts. 2020;

6. Goyal N, Male S, Al Wafai A, Bellamkonda S, Zand R. Cost burden of stroke mimics and transient ischemic attack after intravenous tissue plasminogen activator treatment. Journal of Stroke and Cerebrovascular Diseases. 2015;

7. Ocampo FF, De Leon-Gacrama FRG, Cuanang JR, Navarro JC. Profile of stroke mimics in a tertiary medical center in the Philippines. Neurol Asia. 2021;26(1).

8. Prevention C for disease control and. Stroke Facts. 2020.

9. Warner JJ, Harrington RA, Sacco RL, Elkind MSV. Guidelines for the Early Management of Patients With Acute Ischemic Stroke: 2019 Update to the 2018 Guidelines for the Early Management of Acute Ischemic Stroke. Stroke [Internet]. 2019 Dec 1 [cited 2024 Mar 15];50(12):3331–2. Available from: https://pubmed.ncbi.nlm.nih.gov/31662117/

10. Okano Y, Ishimatsu K, Kato Y, Yamaga J, Kuwahara K, Okumoto K, et al. Clinical features of stroke mimics in the emergency department. Acute Medicine & Surgery. 2018;

11. The National Institute of Neurological Disorders and Stroke rt-PA Stroke Study Group. Tissue Plasminogen Activator for Acute Ischemic Stroke. New England Journal Of Medicine [Internet]. 1995 Dec 14 [cited 2023 Mar 16];333(24):1581–8. Available from: https://www.nejm.org/doi/full/10.1056/nejm199512143332401

12. Zinkstok SM, Engelter ST, Gensicke H, Lyrer PA, Ringleb PA, Artto V, et al. Safety of thrombolysis in stroke mimics: Results from a multicenter cohort study. Stroke. 2013;

13. Matuja SS, Khanbhai K, Mahawish KM, Munseri P. Stroke mimics in patients clinically diagnosed with stroke at a tertiary teaching hospital in Tanzania: A prospective cohort study. BMC Neurol [Internet]. 2020 Jul 7 [cited 2024 Mar 13];20(1):1–7. Available from: https://bmcneurol.biomedcentral.com/articles/10.1186/s12883-020-01853-7

14. Carlos Arturo Sarmiento Limas. ANEXO 3.1 COMENTARIOS A NIVELES DE COMPLEJIDAD Y ACTIVIDADES DE PROMOCIÓN DE LA SALUD Y PREVENCIÓN DE LA ENFERMEDAD [Internet]. 2009 [cited 2024 Mar 16]. Available from: https://www.minsalud.gov.co/Normatividad%20CRES/Acuerdo%2008%20de%202009%20-%20Anexo%203%20-%20Comentarios%20complejidad%20y%20promocion%20de%20la%20salud%20y%20prevencion%20enfermedad.pdf

15. Gottret P, Schieber G. Health Financing Revisited. Health Financing Revisited. 2006 Mar 30;

